# External validation of the predictive performance of population pharmacokinetic models for phenobarbital in pediatric patients

**DOI:** 10.1101/2020.09.10.20192005

**Authors:** Sunae Ryu, Woo Jin Jung, Zheng Jiao, Jung-Woo Chae, Hwi-yeol Yun

**Author notes:** Both of authors should be considered joint first author (Sunae Ryu, Woo Jin Jung). Both of authors should be considered joint corresponding author (Jung-Woo Chae, Hwi-yeol Yun). **Correspondences** Professor Hwi-yeol Yun, PhD., College of Pharmacy, Chungnam National University, Daejeon, Republic of Korea, Professor Jung-woo Chae, PhD., College of Pharmacy, Chungnam National University, Daejeon, Republic of Korea.

## Abstract

**Aim:** Several studies have reported population pharmacokinetic models for phenobarbital (PB), but the predictive performance of these models has not been well documented. This study aims to do external validation of the predictive performance in published pharmacokinetic models.

**Methods:** Therapeutic drug monitoring data collected in neonates and young infants treated with PB for seizure control, was used for external validation. A literature review was conducted through PubMed to identify population pharmacokinetic models. Prediction- and simulation-based diagnostics, and Bayesian forecasting were performed for external validation. The incorporation of size or maturity functions into the published models was also tested for prediction improvement.

**Results:** A total of 79 serum concentrations from 28 subjects were included in the external validation dataset. Seven population pharmacokinetic studies of PB were selected for evaluation. The model by Voller *et al*. [27] showed the best performance concerning prediction-based evaluation. In simulation-based analyses, the normalized prediction distribution error of two models (those of Shellhaas *et al*. [24] and Marsot *et al*. [25]) obeyed a normal distribution. Bayesian forecasting with more than one observation improved predictive capability. Incorporation of both allometric size scaling and maturation function generally enhanced the predictive performance, but with marked improvement for the adult pharmacokinetic model.

**Conclusion:** The predictive performance of published pharmacokinetic models of PB was diverse, and validation may be necessary to extrapolate to different clinical settings. Our findings suggest that Bayesian forecasting improves the predictive capability of individual concentrations for pediatrics.

**What is already known about this subject:**

- Pharmacokinetics of phenobarbital (PB) have been reported large inter-individual difference and treatment with PB required therapeutic drug monitoring.
- Population pharmacokinetic models for PB have been studied by several researchers but predictive performance of these models has not been well documented.

**What this study adds:**

- Predictive performance of pharmacokinetic models of phenobarbital (PB) was various and required validation for extrapolation to different clinical settings.
- Bayesian forecasting could improve the predictability for individual drug concentrations.
- Imputation of both size and maturation functions could help to enhance the predictability of pharmacokinetic models for pediatric patients.

## INTRODUCTION

Phenobarbital (PB) is an antiepileptic drug (AED) targeting the γ-aminobutyric acid type A (GABAA) receptor, which provides a strong inhibitory response in the brain and controls seizures [1]. It is the gold standard treatment for partial onset of seizures and generalized seizures [1–4]. Particularly, it is recommended as the first-line treatment for neonatal seizure, which is a common neurological event in both term and preterm newborn infants and can be associated with neurological dysfunction or adverse cognitive outcomes requiring adequate treatment or management [1, 2, 5].

The half-life of PB is approximately 100 h in adults and 141 h in preterm infants, decreasing to 67 h in infants at 4 weeks old [6]. There is no clear consensus regarding the target therapeutic range of PB [7], but 10–40 mg/L has been reported to be effective and safe [8–13]. Serum concentrations of PB up to 100 mg/L are needed in some infants with refractory seizures [9]. Guidelines for the dosing regimen of PB for pediatric and neonatal patients are available based on body weight, and generally a loading dose of 20 mg/kg with a maintenance dose of 2.5–5 mg/kg/day by slow intravenous injections or by mouth in neonates are suggested [8]. However, optimal dose for neonates is still debating. Calvier *et al*. [14] reported that scaling of the pharmacokinetic parameters based on body weight is reasonable only for patients over 5 years old, and prediction error increases in those below this age. Bettino *et al*. [15] recommended a lower dose per kilogram during the neonatal period due to the longer mean terminal half-life, while other groups have recommended a higher intravenous loading dose of 30 mg/kg/day followed by a maintenance dose of 4–6 mg/kg/day in neonates [16, 17].

Therapeutic drug monitoring (TDM) or clinical pharmacokinetic consultation services (CPCS) are commonly used for seizure care with PB [18], due to the substantial variability of its pharmacokinetic characteristics [15]. Despite individualization of dosing by TDM or CPCS for PB, clinically significant differences between the dose and blood concentration in individual patients have still been reported.

Population pharmacokinetic (popPK) analysis is a practical method for accurate and precise estimation of blood concentrations in pediatric patients [19–22]. The most important issue in modeling approaches is whether the established model can predict the pharmacokinetics in a prospective study or can be extrapolated to different clinical settings, such as patients at another institution, particularly when the purpose of modeling is prediction of optimal individual dose [29]. Recently, numerous studies have evaluated the predictive capability of popPK models [30–37]. Several papers have indicated that the estimation with at least one prior observation using the population information (Bayesian forecasting) could make better predictive capability for individual predictions [30, 36–38].

In this study, we performed external validation of the published popPK models of PB to evaluate their predictive performance based on prediction-based diagnostics, simulation-based diagnostics, and Bayesian forecasting methods. In addition, incorporation of size or maturity functions into the published models was evaluated to improve their predictive capability in neonatal and pediatric populations.

## METHODS

### Data collection for external validation

External validation of PB was performed using the dataset of Back *et al*. [39] utilizing TDM data of pediatrics in the neonatal intensive care unit (NICU) of Kyunghee University hospital in patients receiving treatment with PB for seizure control. In that study, PB was administered in accordance with the general guidelines at an initial dose of 15–20 mg/kg followed by a maintenance dose of 3– 5 mg/kg/day from 12–24 h after the initial dose [8, 43]. Blood samples were taken between 5 min and 3 h before the subsequent dose and drug concentrations were analyzed using a turbidimetric immunoassay (COBAS 6000; Roche, Grenzach-Wyhlen, Germany). Clinical and demographic records of the patients were collected to cover covariates in published popPK models. Gestational age (GA), postnatal age (PNA), postconceptional age (PCA), height, weight, body mass index (BMI), and weight at birth were recorded. Laboratory test results, such as serum levels of creatinine, aspartate aminotransferase (AST), alanine aminotransferase (ALT), total bilirubin, urine output, and Apgar scores at 1 min and 5 min, were extracted. Fat-free mass (FFM) was calculated as described previously [44]. In the case of missing covariate information for SCR, the next value observed in the subject was carried backward. Approval for this study was provided by the institutional ethics committee of Kyunghee University Hospital (IRB file no. 2015–01–026–002, May 29. 2016).

### Selection of published popPK models

A literature review was conducted through PubMed to screen for all published papers describing popPKs of PB between January 1980 and December 2019. The search was performed using the terms (“phenobarbital”) AND (“population”) in the abstract field. After scrutinizing the abstracts, reports were included if they developed popPK models of PB reporting pharmacokinetic model parameters and were written in English. Studies in patients of any age group, including adults, were included as incorporation of the size and maturation functions into the reported model would be performed. Reviews or studies with overlapping data or external validation of another model were excluded.

### External validation of predictive capability of popPK models

Published pharmacokinetic models for external validation were regenerated by nonlinear mixed effects modeling using NONMEM 7.4 (Icon Development Solutions, Ellicott City, MD) assisted by Per-Speaks-NONMEM (PsN) 4.8.1 (https://uupharmacometrics.github.io/PsN/) and Pirana 2.9.9 (http://www.pirana-software.com). Each popPK model was reconstructed as described in the literature, and the parameters were set to the published values. The NONMEM subroutine ADVAN6 was used for all candidate models as the external dataset contained data from both intravenous (IV) and oral administration. For models that used only IV data, and where the first-order absorption constant (*Ka*) was not reported, it was arbitrarily fixed at 50 h^−1^. For the next step, individual serum concentrations of PB were predicted for each time point in the validation dataset based on the reconstructed popPK models on a one by one basis. Prediction-based diagnostics, simulation-based diagnostics, and Bayesian forecasting were performed to evaluate the predictive capability of the candidate models.

### Prediction-based diagnostics

Predictive performance was evaluated as the bias and precision using the prediction error percentage (PE[%]), mean prediction error (MPE), median absolute prediction error (MAPE), and root mean squared prediction error (RMSE) [45]. The population predicted concentration (*Cpred*) of the external dataset was estimated by simulation of the reconstructed models with reported parameter estimates and compared to corresponding observed concentrations (*Cobs*). PE(%), MPE,

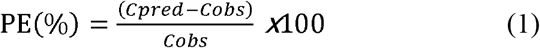

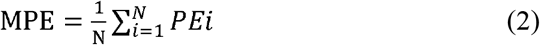

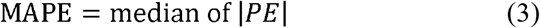

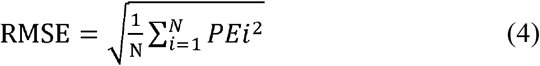

The PE(%) values that fell within the range of 20% (F_20_) and 30% (F_30_) were compared as proposed previously [36].

### Simulation-based diagnostics

The normalized prediction distribution error (NPDE) was computed using the add-on R package, NPDE (https://CRAN.R-project.org/package = NPDE), in R 3.5.1 software (https://www.rproject.org) and R studio 1.1 (www.rstudio.com) [46]. Simulated data files were generated using NONMEM with 1000 simulations with the fixed final parameters in the reported models. When the reported model adequately describes the external validation dataset, NPDE will follow a standard normal distribution, with a mean of zero and variance of 1. The assumption of *N* (0, 1) distribution was tested using Wilcoxon’s signed rank test for means, Fisher’s variance test, and Shapiro–Wilks test for normality. To visualize the distribution, quantile-quantile (Q-Q) plots, histograms of NPDE, scatter plots of NPDE versus each time point, or predicted concentrations were plotted in a predefined output format provided by the NPDE package in R.

### Bayesian forecasting

Bayesian estimation with maximum a posteriori estimation was performed to investigate whether predictive capability of the popPK models on the external dataset could be improved by updating the model with observed concentrations [47]. Fixed and random effects were set to the reported final values in the literature, but individual pharmacokinetic parameters were updated based on the first one, two, three, or all prior measurements for all subjects via first-order (FO) estimation with the POSTHOC option in NONMEM. Then, individual predicted concentrations (*C_ipred_*) were obtained for all of the measured time points and compared to the corresponding observed concentrations (*C_obs_*) to calculate the individual prediction error (IPE, Equation 5).

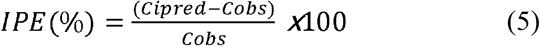

### Incorporation of size and maturation function

The influences of the size and maturation function on neonates and infants were assessed by incorporating them into the candidate models if they did not originally include these terms. Exponents of 1 and 0.75 were used for allometric scaling on the body weight to reflect the size for clearance (*CL*) and volume of distribution (*V_d_)*, respectively (Equation 6). A sigmoidal *E_max_* function on PCA was applied to the *CL* with the reported Hill coefficient and maturation half-life (*TM_50_*) as suggested by Back *et al*. [39] (Equation 7).

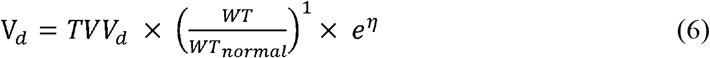

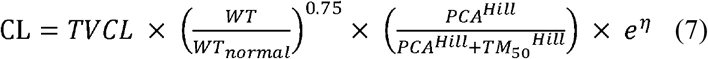

## RESULTS

### External validation dataset

A total of 79 trough serum concentrations from 28 neonates were included in this study. A summary of the demographic and clinical characteristics of the patients is presented in Table 1. The mean postnatal age was 32.4 days, ranging from 3 to 150 days. The mean body weight was 3.3 kg (1–6.9 kg). The mean daily dose of PB was 1.08 mg/kg/day (0.21–4.24 mg/kg/day) given either intravenously or orally. The mean serum concentration was 20.3 mg/L (5.3–42.3 mg/L) with a median sample time of 11.5 h after the previous dose.

**Table 1.** Demographic characteristics of external validation dataset

| Characteristics | Mean ± SDs | Range |
| --- | --- | --- |
| No. of patients (preterm infants) | 28 (11) |  |
| No. of concentrations | 79 |  |
| Gender |  |  |
| Male/Female | 11/17 |  |
| Age |  |  |
| GA (weeks) | 36.7 ± 4.4 | 23.6–41.7 |
| PNA (days) | 32.4 ± 30.9 | 3–150 |
| PCA (weeks) | 38.6 ± 3.3 | 31–51.1 |
| Body weight (kg) | 3.3 ± 1 | 1–6.9 |
| Birth weight (kg) | 2.64 ± 0.87 | 0.4–3.81 |
| Height (cm) | 50.6 ± 5.8 | 31–63.2 |
| Daily dose (mg/kg/day) | 1.08 ± 0.61 | 0.21–4.24 |
| Serum concentration (mg/L) | 20.3 ± 9.2 | 5.3–42.3 |
| Sampling time (hours)<sup>a</sup> | 11.5 | 7.5–24 |
| Time to last sampling (days) | 26.8 ± 29.0 | 1.5–99.0 |
| 5-min Apgar score | 4.8 ± 3.3 | 1–10 |

GA, gestational age; PNA, postnatal age; PCA, post-conceptional age; SD, standard deviation;
<sup>a</sup> sampling time was expressed in a median time after dose

### Review of published popPK models

Among 564 articles with the terms “phenobarbital” and “population” appearing in their summaries published between 1980 and 2019, 10 dealt with modeling or popPK analyses of PB. A detailed flow chart of inclusion and exclusion of papers is shown Figure S1. After processing to select suitable articles, a total of seven studies with nonlinear mixed effect models were included in this study. Grasela *et al*. [28] was performed in 1985 and others were reported between 2011 and 2018. Three studies were conducted in the USA [17, 24, 28], three were conducted in Europe (the Netherlands [27], Serbia [26], and France [25]), and one was conducted in Japan [23]. One model was developed with data from adults [26] and one was from pediatric patients < 19 years old [17]. The remaining five models were established with data from neonates or infants [23–25, 27, 28]. Only two studies were conducted with sample sizes of more than 100 patients [17, 26]. The demographic and pharmacokinetic characteristics of the published models are summarized in Table 2.

**Table 2.** Demographic characteristics of population PK models in published papers

| Study (Year) | Country | No. of Patients/<br>Observations | Subjects<br>(Age) | Dosing | Concentrations(mg/L) | Assay Method |
| --- | --- | --- | --- | --- | --- | --- |
| Moffett et al<br>(2018) | USA | 355/NA | Pediatric, <19 years<br>(14.6 weeks,<br>IQR[3.1, 42.8]) | LD 10-20mg/kg<br>MD 3-8mg/kg | 41.1±23.9 | Chemiluminescent<br>microparticle immunoassay<br>(VITROS) |
| Voller et al<br>(2017) | Netherlands | 53/229 | Neonates<br>(4.5days, [0-22]) | LD 20mg/kg(4-40.7)<br>MD 3.9 mg/kg (1.3-20) | Range 3.2-75.2 | Fluorescence polarization<br>immunoassay (COBAS<br>INTEGRA) |
| Vucicevic et al<br>(2015) | Serbia | 136/205 | Adults<br>(42.04 years, SD 13) | 130.2±58.37mg/day | 19.26±9.003 | Homogeneous enzyme<br>immunoassay (Emit 2000) |
| Marsot et al<br>(2014) | France | 48/94 | Neonates and infants<br>(3.8weeks, [0-29.4]) | LD 10-20mg/kg<br>MD 5mg/kg | 26±9.8 | Immunoassay (Microgenic<br>reagents, Olympus AU400) |
| Shellhaas et al<br>(2013) | USA | 39/164 | Term neonates with<br>HIE<br>(13.1days) | NA | NA | NA |
| Yukawa et al<br>(2011) | Japan | 70/109 | Neonates and infants<br>(15.8days [1-73]) | 23.5±16.3<br>mg(suppository)<br>7.8±2.1 mg(powder) | 29.7±19.0 (suppository)<br>10.7±4.6 (powder) | Immunoassay (Emit 2000<br>Phenobarbital assay reagent,<br>COBAS-FARA) |
| Grasela et al<br>(1985) | USA | 59/160 | Preterm infants<br>(8.2days [1-16]) | NA | NA | HPLC |

NA, not available; IQR, interquartile range; IV, intravenous; HIE, hypoxic ischemic encephalopathy; LD, loading dose; MD, maintenance dose; HPCL, high-pressure liquid chromatography; Ages are expressed either in median or mean postnatal age with range in bracket if not specified; Concentrations are expressed in mean ± standard deviation (SD) if not specified.

All of the studies were fitted using a one-compartment model. Five had both IV and oral administration data, and estimations were performed using the ADVAN2 TRANS2 subroutine in NONMEM. Two other models were constructed with only IV data, for which the absorption rate constant (*K_a_*) was fixed to an arbitrary value of 50. In the remaining models that reported *K_a_* or bioavailability (*F*), the reported values in the literature were used for external validation. Interindividual variability was described with an exponential model [17, 24–27] or in proportional error model [23, 28]. The residual variation was described with a proportional model [17, 23, 26– 28], additive model [25], or a combined proportional and additive model [24].

All models that were constructed with pediatric patients indicated that body size was a predictor of both *CL* and *V_d_*. Meanwhile, body weight was not relevant in the model for adult subjects [26]. As a maturation factor, PNA or PMA was incorporated into four models to reflect physiological development in neonates [17, 23, 24, 27]. The detailed information for the popPK models in each published paper is summarized in Table 3.

**Table 3.**
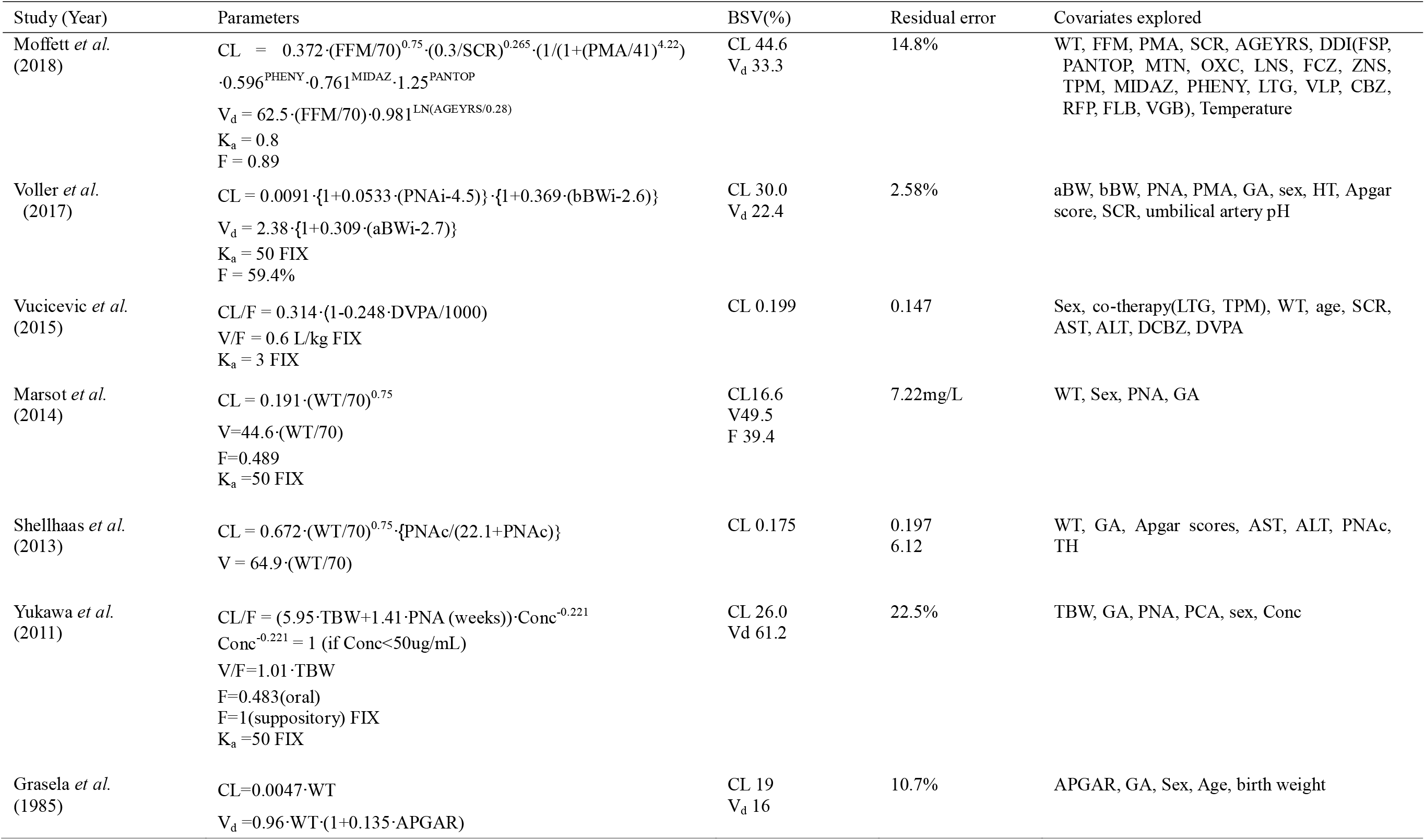
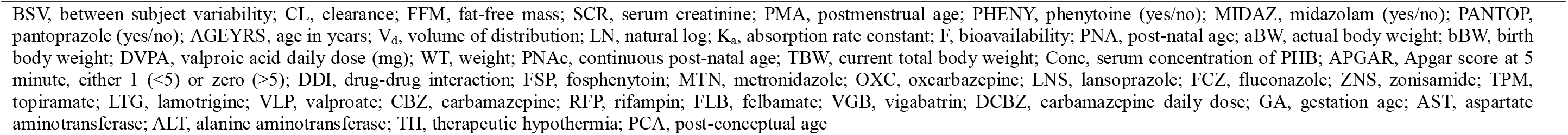
Population PK models in published papers

| Study (Year) | Parameters | BSV(%) | Residual error | Covariates explored |
| --- | --- | --- | --- | --- |
| Moffett *et al.* (2018) | $CL = 0.372 \cdot (FFM/70)^{0.75} \cdot (0.3/SCR)^{0.265} \cdot (1/(1+(PMA/41)^{4.22})) \cdot 0.596^{PHENY} \cdot 0.761^{MIDAZ} \cdot 1.25^{PANTOP}$<br>$V_d = 62.5 \cdot (FFM/70) \cdot 0.981^{LN(AGEYRS/0.28)}$<br>$K_a = 0.8$<br>$F = 0.89$ | CL 44.6<br>V<sub>d</sub> 33.3 | 14.8% | WT, FFM, PMA, SCR, AGEYRS, DDI(FSP, PANTOP, MTN, OXC, LNS, FCZ, ZNS, TPM, MIDAZ, PHENY, LTG, VLP, CBZ, RFP, FLB, VGB), Temperature |
| Voller *et al.* (2017) | $CL = 0.0091 \cdot \{1 + 0.0533 \cdot (PNAi - 4.5)\} \cdot \{1 + 0.369 \cdot (bBW_i - 2.6)\}$<br>$V_d = 2.38 \cdot \{1 + 0.309 \cdot (aBW_i - 2.7)\}$<br>$K_a = 50$ FIX<br>$F = 59.4\%$ | CL 30.0<br>V<sub>d</sub> 22.4 | 2.58% | aBW, bBW, PNA, PMA, GA, sex, HT, Apgar score, SCR, umbilical artery pH |
| Vucicevic *et al.* (2015) | $CL/F = 0.314 \cdot (1 - 0.248 \cdot DVPA/1000)$<br>$V/F = 0.6$ L/kg FIX<br>$K_a = 3$ FIX | CL 0.199 | 0.147 | Sex, co-therapy(LTG, TPM), WT, age, SCR, AST, ALT, DCBZ, DVPA |
| Marsot *et al.* (2014) | $CL = 0.191 \cdot (WT/70)^{0.75}$<br>$V = 44.6 \cdot (WT/70)$<br>$F = 0.489$<br>$K_a = 50$ FIX | CL 16.6<br>V 49.5<br>F 39.4 | 7.22mg/L | WT, Sex, PNA, GA |
| Shellhaas *et al.* (2013) | $CL = 0.672 \cdot (WT/70)^{0.75} \cdot \{PNAc/(22.1 + PNAc)\}$<br>$V = 64.9 \cdot (WT/70)$ | CL 0.175 | 0.197<br>6.12 | WT, GA, Apgar scores, AST, ALT, PNAc, TH |
| Yukawa *et al.* (2011) | $CL/F = (5.95 \cdot TBW + 1.41 \cdot PNA(\text{weeks})) \cdot Conc^{-0.221}$<br>$Conc^{-0.221} = 1$ (if $Conc < 50 \mu\text{g/mL}$)<br>$V/F = 1.01 \cdot TBW$<br>$F = 0.483$(oral)<br>$F = 1$(suppository) FIX<br>$K_a = 50$ FIX | CL 26.0<br>V<sub>d</sub> 61.2 | 22.5% | TBW, GA, PNA, PCA, sex, Conc |
| Grasela *et al.* (1985) | $CL = 0.0047 \cdot WT$<br>$V_d = 0.96 \cdot WT \cdot (1 + 0.135 \cdot APGAR)$ | CL 19<br>V<sub>d</sub> 16 | 10.7% | APGAR, GA, Sex, Age, birth weight |

BSV, between subject variability; CL, clearance; FFM, fat-free mass; SCR, serum creatinine; PMA, postmenstrual age; PHENY, phenytoine (yes/no); MIDAZ, midazolam (yes/no); PANTOP, pantoprazole (yes/no); AGEYRS, age in years; $V_d$ , volume of distribution; LN, natural log; $K_a$ , absorption rate constant; F, bioavailability; PNA, post-natal age; aBW, actual body weight; bBW, birth body weight; DVPA, valproic acid daily dose (mg); WT, weight; PNAc, continuous post-natal age; TBW, current total body weight; Conc, serum concentration of PHB; APGAR, Apgar score at 5 minute, either 1 ( $<5$ ) or zero ( $\geq 5$ ); DDI, drug-drug interaction; FSP, fosphenytoin; MTN, metronidazole; OXC, oxcarbazepine; LNS, lansoprazole; FCZ, fluconazole; ZNS, zonisamide; TPM, topiramate; LTG, lamotrigine; VLP, valproate; CBZ, carbamazepine; RFP, rifampin; FLB, felbamate; VGB, vigabatrin; DCBZ, carbamazepine daily dose; GA, gestation age; AST, aspartate aminotransferase; ALT, alanine aminotransferase; TH, therapeutic hypothermia; PCA, post-conceptual age

### Prediction-based diagnostics

Diagnostic plots of the population predictions of each model with fixed final parameters versus measured concentrations of the external validation dataset showed that the model results of Voller *et al*. [27] were considerably well aligned with the unit line (*y* = *x*, red line in Figure S2). Meanwhile, those of Shellhaas *et al*. [24] and Yukawa *et al*. [23] were underestimated. The model of Vucicevic *et al*. [26], which was constructed for an adult population, showed a skewed distribution due to differences in age from our external validation dataset (Figure S2).

To quantify predictive performance, MPE and MAPE were calculated for bias and precision, respectively [45]. The model proposed by Marsot *et al*. [25] showed superior accuracy (MPE 0.17%) followed by that of Voller *et al*. [27] (MPE –2.61%). For precision, the model of Voller *et al*. [27] showed the best MAPE of 29.86% followed by those of Shellhaas *et al*. [24] and Yukawa *et al*. [23] with MAPEs of 32.08% and 32.32%, respectively (Figure 1, Table S1). The model of Vucicevic *et al*. [26] is not shown in Figure 1 because of the skewed distribution.

**Figure 1.**
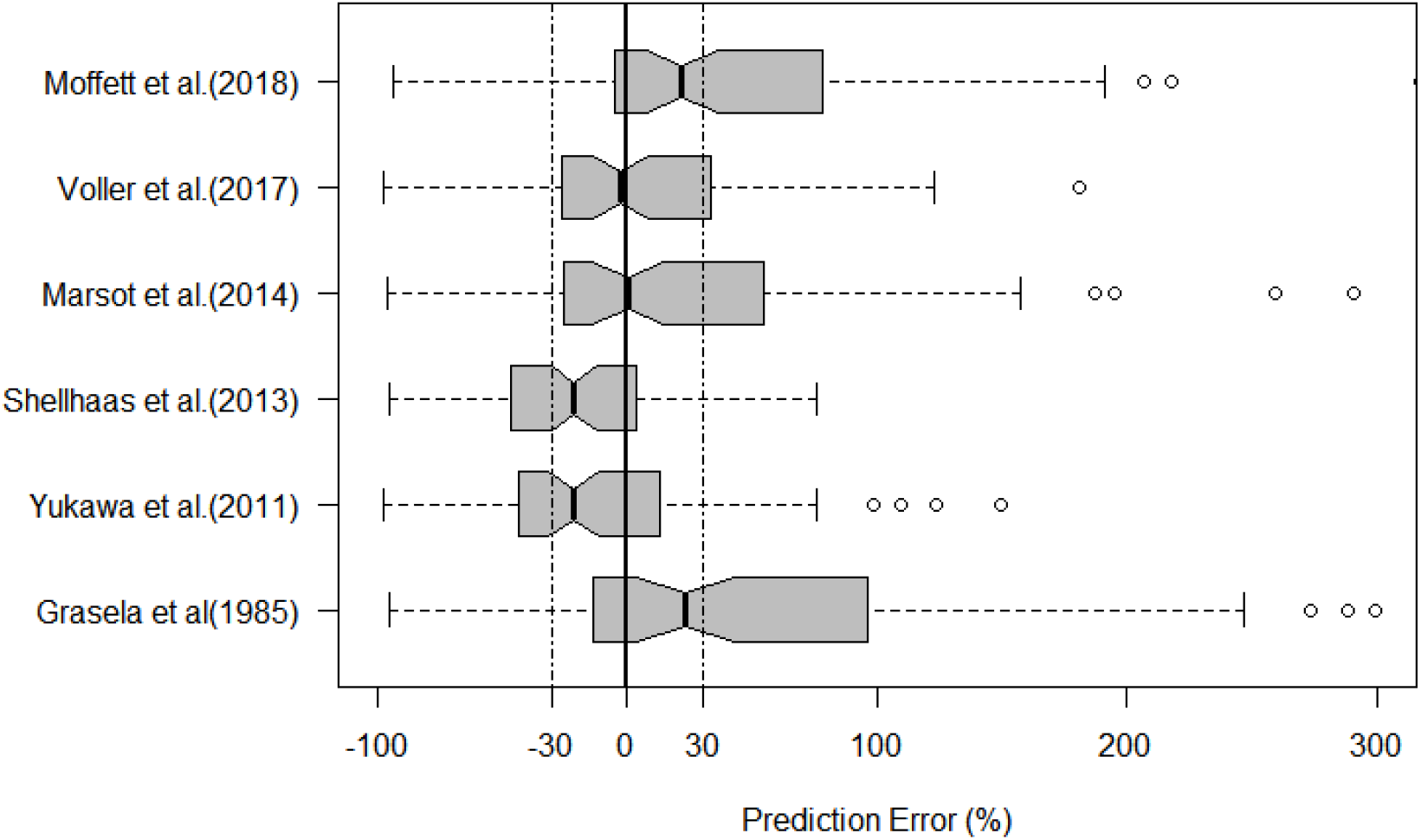
Box plots of prediction errors of population PK models of phenobarbital. Thick solid line represents the mean error of zero and dot dashed line corresponds to the range of -30% and 30% difference from mean error.

The percentages of PE falling into the range of 20% (F_20_) or 30% (F_30_) were calculated. The model proposed by Voller *et al*. [27] was superior to the others with an F_20_ of 35.44% and F_30_ of 50.63%. Among the others, only the model of Moffett *et al*. [17] showed F_20_ > 30%, and four models (those of Shellhaas *et al*. [24], Yukawa *et al*. [23], Marsot *et al*. [25], and Moffett *et al*. [17]) showed F_30_ > 40% (Table S1). Overall, the model of Voller *et al*. [27] showed the best performance in terms of prediction-based evaluation.

### Simulation-based diagnostics

The NPDEs of the models of Marsot *et al*. [25] and Shellhaas *et al*. [24] obeyed a normal distribution with a global test *p* > 0.01 (Figure 2, Table S2). However, those of the others, including Voller *et al*. [27], which showed the best predictive performance in the prediction-based diagnosis, did not (Figure S3).

**Figure 2.**
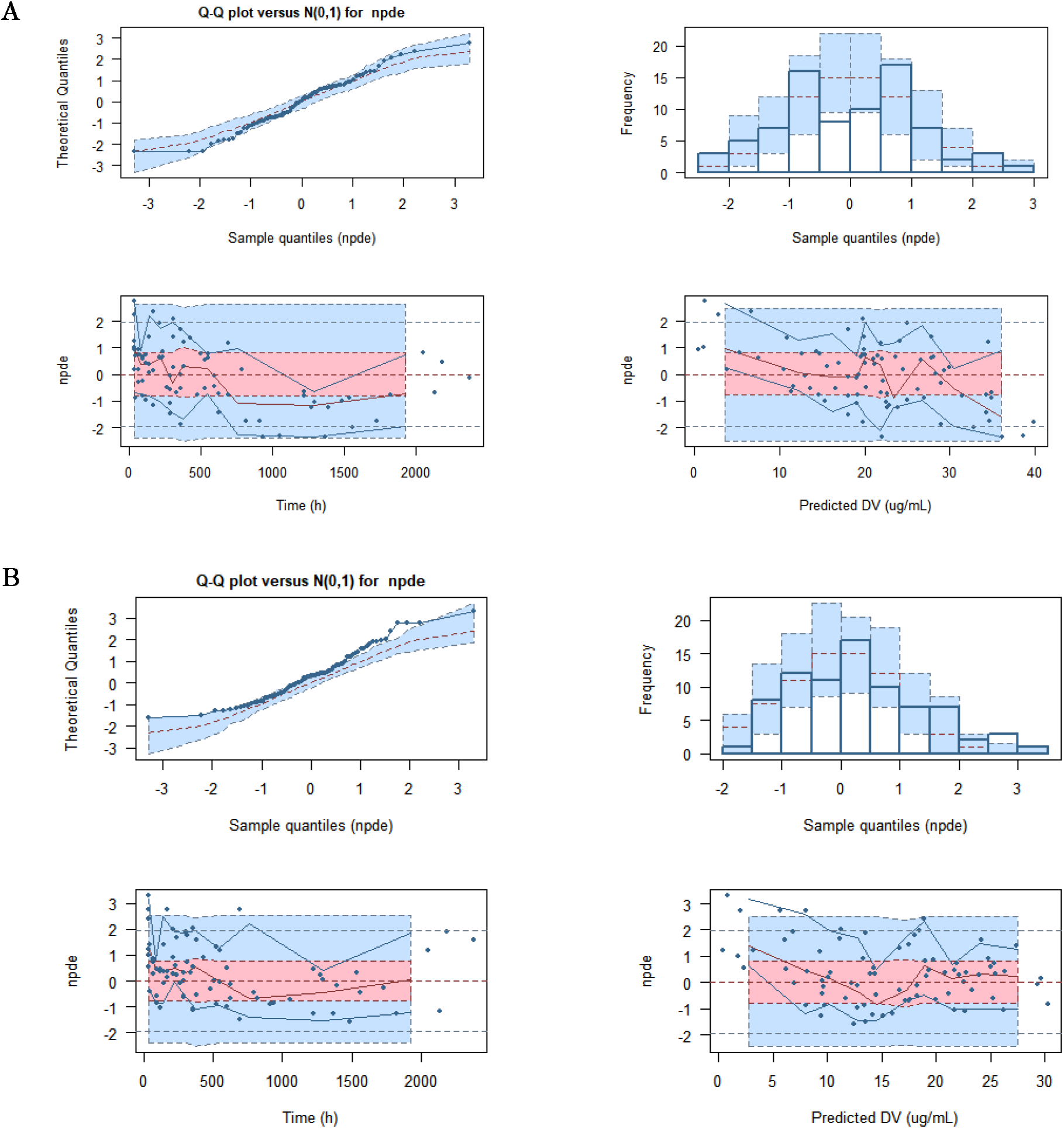
Normalized prediction distribution error (NPDE) plots of external validation data set for the models by Marsot *et al*. (A) and Shellhaas *et al*.(B): Quantile-quantile (q-q) plot of NPDE versus the expected standard normal distribution (upper left). Histogram of npde with the density of the standard normal distribution (upper right). The blue prediction intervals are obtained from the theoretical normal distribution. Scatterplot of the npde versus time after first dose in hour (lower left). The pink areas are the prediction interval for the median, while the blue areas show the prediction areas for the boundaries of the 95% prediction intervals. Scatterplot of npde versus predicted concentrations in ug/mL (lower right).

### Bayesian forecasting

Bayesian forecasting demonstrated that the individual predictive capability was improved by the information of more than one prior measurement in most of the models (Figure 3). Two or three prior observations did not result in further significant improvements except in the models of Marsot *et al*. [25] and Grasela *et al*. [28], where the improvement was obvious with more than three prior samples. The individual predictions for the pediatric population even in the adult model were markedly improved by Bayesian estimation, as for the model of Vucicevic *et al*. [26] (Figure S4).

**Figure 3.**
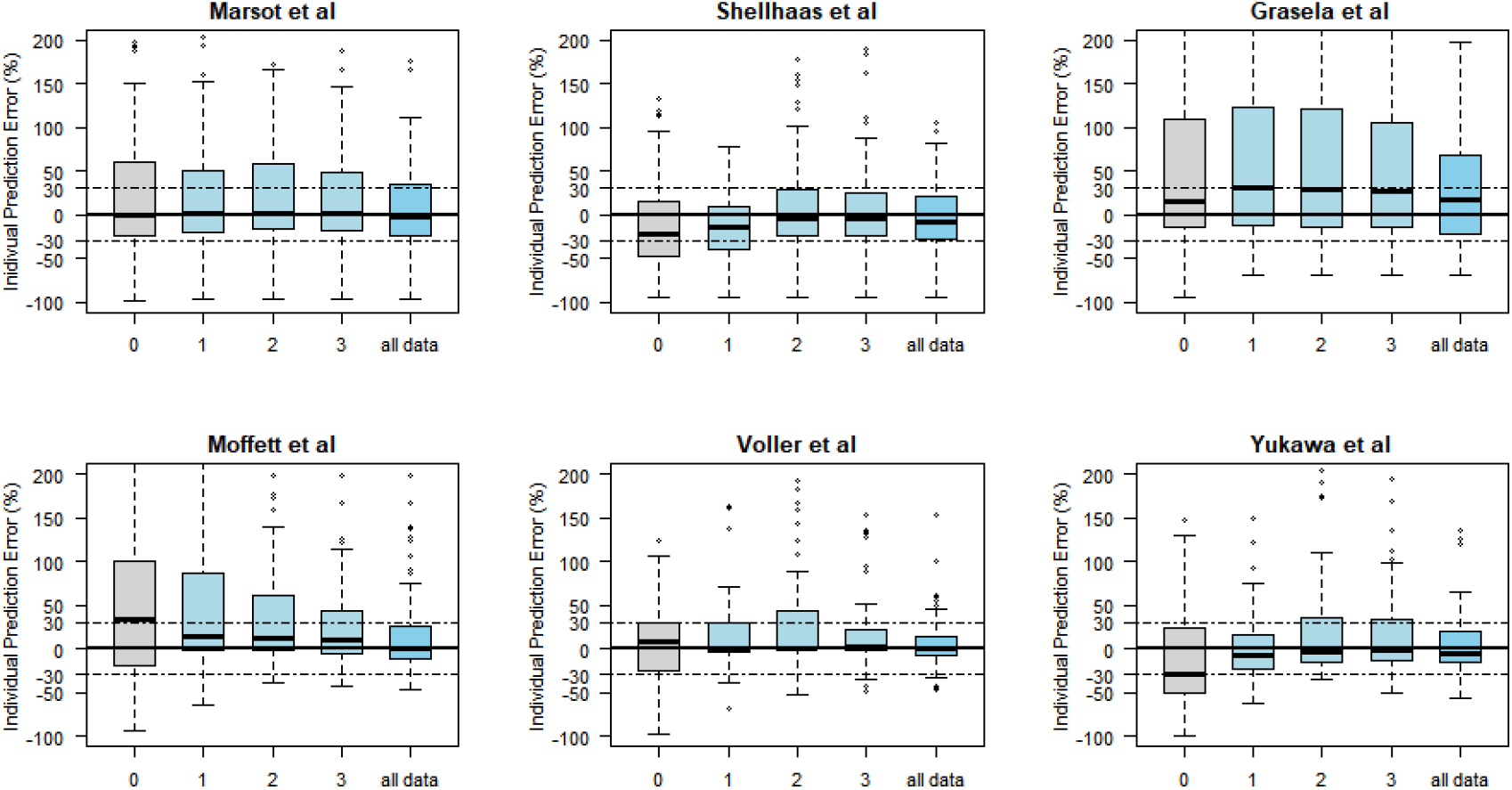
Box plots of individual prediction errors of population pharmacokinetic models of phenobarbital with Bayesian forecasting in different number of prior measurements; 0, 1 2, 3 and all the observed measurements. Thick solid line represents the mean error of 0 and dotdashed line corresponds to the range of -30% and 30%. For the Bayesian estimation of the model by Yukawa *et al*., 5 subjects with negative individual PK parameters estimated were excluded from the analysis.

### Size and maturation function

All of the published models were tested for the effects of allometric size and sigmoidal maturation function to improve predictive capability for pediatric populations. As three studies already included allometric size scaling in the original models, only the maturation function was tested as an added function [17, 24, 25]. Allometric size scaling and the sigmoidal maturation function were substituted for the originally imputed size and maturation effect in the remaining models.

The model of Vucicevic *et al*. [26] was markedly improved by the incorporation of allometric size function as this study was performed with adult patients only (Figure 4). The influence of the application of maturity factors varied: one improved significantly [25], two improved marginally [24, 28], but three models showed slight worsening [17, 23, 27] (Figure S5). Generally, four of the seven models investigated showed improvement with the incorporation of size and maturation function [24–26, 28].

**Figure 4.**
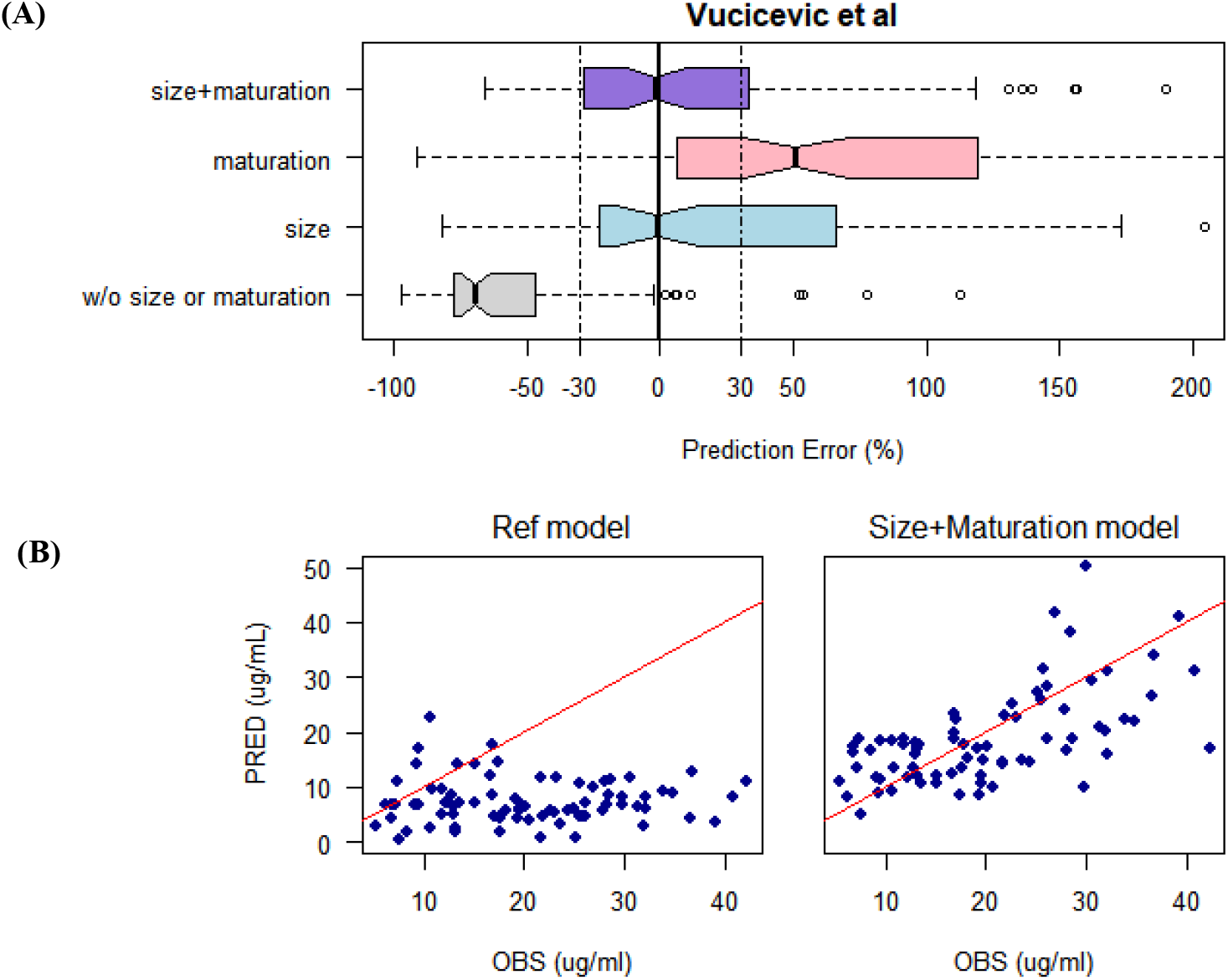
(A) Box plots of prediction errors without or with incorporation of size or maturation function of the model by Vucicevic *et al*. Box plots were plotted from the predicted concentrations with estimated thetas with the external dataset, without size or maturation function, with size function, with maturation function, or with both size and maturation function, respectively. Thick solid line represents the mean error of 0 and dotdashed line corresponds to the range of -30% and 30%. (B) Diagnostic plots of predicted versus observed measurement before (left) and after (right) incorporation of size or maturation function into the model by Vucicevic *et al*. The led line represents the identity line.

## DISCUSSION

External validation was performed for seven published popPK models of PB to evaluate their predictive performance using an independent TDM dataset from 28 neonates, including 11 preterm infants. Those studies validated the models using internal and/or external validation methods. Internal validation using diagnostic plots, including predicted concentrations versus observations, was performed most frequently (6 of 7 papers). Bootstrap techniques comparing the final parameter estimates with their corresponding bootstrap estimates and 95% confidence intervals of 1000 replicates of the dataset were also used frequently (5 studies). As advanced internal validation, NPDE, prediction- and variability-corrected visual predictive check (pvcVPC), or predictive performance check via mean prediction error (ME) or mean absolute prediction error (MAE) were also performed. Grasela *et al*. [28] and Voller *et al*. [27] included external validation with a separate dataset that was not used in building the popPK models. Considering that a systematic review indicated that 45% of the pharmacokinetic models from all population models published between 2002 and 2004 were only subjected to basic internal validation (GOF) and not more than 7% were subjected to external validation [29], the validation method involved in the candidate models was assumed to be comparably good.

In the prediction-based diagnostics, the model of Voller *et al*. [27] showed the best predictive capability. Voller *et al*. [27] studied 53 preterm or term neonates in a TDM setting to build a popPK model and performed external validation with 17 preterm neonates from an ongoing prospective study. They reported an MPE of –8.4% in their external validation; our results are comparable (5.98%). In the present study, the precision (RMSE) was 8.2 μg/mL, which was the best figure among the candidate models. In addition, F_20_ and F_30_ were superior to the other models (35% and 51%, respectively). Two external validation studies set the acceptability criteria to 35% and 50% for F_20_ and F_30_, respectively [36, 37]. This was the only model that met these criteria and showed the best predictive capability determined according to the prediction-based evaluation. This is reasonable as the demographic characteristics of the study population, treatment period, loading/maintenance dosages, and bioassay method were comparable to those of the external validation dataset. Grasela *et al*. [28] reported an MPE and RMSE of 1.0 μg/mL and 5.2 μg/mL, respectively, for a model that included its own external validation with 15 additional patients. In the present study, these values were 5.05 μg/mL and 13.17 μg/mL, respectively. The considerable heterogeneity in the population groups between the two studies could account for these discrepancies and poor predictive capability. The average study periods in the literature are 8.2 days (range 1–16 days) and 7.3 days (range 4–9 days) for the study group and the validation group, respectively, while it was 26.8 days (range 1.9–99.0 days) in our data. There were also large differences in body weight and GA of the study populations. In addition, the serum concentration was measured using HPLC in previous studies, while we used an immunoassay.

In the simulation-based evaluation, the models of Marsot *et al*. [25] and Shellhaas *et al*. [24] described the external validation dataset adequately, accepting the null hypothesis that the NPDE followed the normal distribution. Although it had the best predictive capability in prediction-based diagnostics, the NPDE of the model by Voller *et al*. [27] did not follow a normal distribution. Inconsistencies between prediction-based evaluation and simulation-based evaluation have also been reported by other groups [36, 37]. As noted by Karlsson and Sevic [48], the generation of appropriate simulations for NPDE analyses could be an issue in the TDM setting. The treatment period for our dataset ranged from 1.9 days to 99.0 days, and the sampling number varied from 1 to 8. This implies that there was considerable heterogeneity in the dose–pharmacokinetic–response relations between subjects and may have resulted in inadequate simulations for NPDE evaluation.

In Bayesian forecasting, the individual predictive capability was improved in most of the models with at least one prior observation. This result is reasonable, as the benefits of Bayesian forecasting are well documented [30, 36–38, 49]. Meanwhile, the model of Marsot *et al*. [25], which showed comparable predictive capability without posterior information, was not markedly improved. The variability in individual prediction error was increased after adding posterior observations in the model by Grasela *et al*. [28]. As expected, the model of Vucicevic *et al*. [26] was markedly improved by incorporating allometric size scaling on *CL* and *V_d_* because the model was built with adult data, and the pharmacokinetic parameters of *CL* and *V_d_* for adults are related to allometric size scaling when applied to pediatric populations. The improvement of predictive capability was further driven by incorporation of PCA in the Michaelis–Menten (MM) equation on *CL* as an index of maturation, consistent with Back *et al*. [39].

Among the other models for pediatric populations, two were improved by the addition of size or maturation factors. Adding maturation factor in the model of Marsot *et al*. [25] rendered a slight improvement by decreasing MPE and MAPE from 14.8% to 3.6% and from 50.6% to 36.3%, respectively. The predictive capability of the model of Grasela *et al*. [28] improved with the inclusion of the size and maturation functions although the authors found no effects of GA on *CL*. They explained that no influence on clearance was detected as their subjects had GA < 34 weeks.

Yukawa *et al*. [23] and Shellhaas *et al*. [24] identified PNA as an important maturation marker, Moffett *et al*. [17] reported PMA as a marker of maturity and Voller *et al*. [27] suggested that PNA and birthweight were correlated with *CL*. For these four models, substitution of the reported covariates with PCA based on the MM equation did not yield benefit with regard to predictive capability. Our results suggest that adding the maturation factor into pharmacokinetic models for pediatric populations could be beneficial for predictive performance, but it was not necessary to substitute PCA based on MM equation in place of the other maturation covariates.

The methodology used for Bayesian forecasting in our study could have limitations. In the present study, all 28 subjects were included in the Bayesian estimation step and each subject contributed one, two, or three observations as long as available. Based on the updated individual pharmacokinetic parameters through Bayesian estimation, the individual predictions were obtained for all time points for the subjects. In studies that have reported successful Bayesian forecasting, only the last observations had been predicted for subjects who met the criterion for the certain number of prior observations [30, 36, 37]. Nonetheless, our results suggest that even a less parsimonious Bayesian forecasting methodology could be useful to predict more precise individual concentrations. The absence of history regarding concomitant drugs in our data may represent another limitation. There is potential for multiple medications during PB therapy [6]. Since PB is largely metabolized by CYP2C9 isozymes and can also act as an inducer of CYP3A4, it shows drug–drug interactions with other CYP substrates [6, 17]. Among the studies included in the investigation, two reported significant drug–drug interactions with valproic acid, phenytoin, midazolam, or pantoprazole [17, 26]. Due to the absence of information on concomitant drugs in our study, the influence of drug–drug interactions could not be examined as a factor affecting the predictive performance of corresponding studies, which may have resulted in misspecification to some extent.

## CONCLUSIONS

Published popPK models of PB showed a wide degree of variation in predictive performance, and validation may be necessary for extrapolation to different clinical settings. The model of Voller *et al*. [27] showed the best performance from the viewpoint of prediction-based evaluation with considerable accuracy and precision. Our findings suggest that Bayesian forecasting could be useful to improve the predictive capability of individual concentrations for pediatric populations. In addition, incorporation of both size and maturation function could help to enhance the predictive performance of PK models for pediatric patients.

## Data Availability

The data that support the findings of this study are available from the corresponding author upon reasonable request.

## ACKNOWLEDGEMENTS

This study was supported by research funds from Chungnam National University and also supported by Institute of Information & communications Technology Planning & Evaluation (IITP) grant funded by the Korea government (MSIT) (No.2020-0-01441, Artificial Intelligence Convergence Research Center (Chungnam National University))

## AUTHOR CONTRIBUTIONS

S.R., J.-W.C., and H.-y.Y. designed and supervised the study. S.R., W.J.J., J.-W.C., and H.-y.Y. performed the experiments. S.R., W.J.J., J.-W.C., and H.-y.Y. analyzed the data. Z.J. provided critical feedback. S.R. and H.-y.Y. wrote the paper with input from all authors

---

**Table S1.** Results of prediction based diagnostics

| Model | MPE<br>(ug/mL) | MAPE<br>(ug/mL) | RMSE<br>(ug/mL) | MPE(%) | MAPE(%) | F<sub>20</sub>% | F<sub>30</sub>% |
| --- | --- | --- | --- | --- | --- | --- | --- |
| Moffett et al | 5.24 | 9.16 | 11.82 | 21.9 | 39.76 | 31.65 | 40.51 |
| Voller et al al | -0.49 | 6.56 | 8.20 | -2.61 | 29.86 | 35.44 | 50.63 |
| Vucicevic et al<br>al | -19.57 | 19.57 | 21.63 | -95.08 | 95.08 | 0 | 0 |
| Marsot et al | 0.91 | 8.17 | 10.16 | 0.17 | 36.42 | 29.11 | 44.30 |
| Shellhaas et al | -5.11 | 7.21 | 9.35 | -21.19 | 32.08 | 25.32 | 46.84 |
| Yukawa et al | -4.45 | 7.55 | 9.38 | -21.55 | 32.32 | 27.85 | 45.57 |
| Grasela et al | 5.05 | 10.57 | 13.17 | 23.47 | 35.92 | 25.32 | 37.97 |

MPE, mean prediction error; MAPE, median absolute prediction error; RMSE, root mean squared error; MPE(%), Median percentage prediction error; MAPE(%), Median absolute percentage error; F<sub>20</sub>%, percentage of prediction error that falls within $\pm 20\%$ ; F<sub>30</sub>%, percentage of prediction error that falls within $\pm 30\%$ .

**Table S2.** Results of simulation based diagnostics using normalized prediction distribution errors, NPDE

| Model | Mean |  | Variance |  | Skewness |  | Kurtosis |  |
| --- | --- | --- | --- | --- | --- | --- | --- | --- |
|  | value | P value<sup>*</sup> | value | P value<sup>†</sup> | value | P value<sup>‡</sup> | value | P value<sup>¶</sup> |
| Moffett et al | -0.54 | 0.004 | 2.59 | <0.001 | 0.39 | 0.066 | -0.09 | <0.001 |
| Voller et al | -0.42 | 0.126 | 5.93 | <0.001 | 0.20 | <0.001 | -1.35 | <0.001 |
| Marsot et al | -0.03 | 0.836 | 1.34 | 0.048 | 0.04 | 0.598 | -0.46 | 0.145 |
| Shellhaas et al | 0.34 | 0.009 | 1.25 | 0.139 | 0.50 | 0.06 | -0.28 | 0.028 |
| Yukawa et al | 0.42 | 0.006 | 1.73 | <0.001 | 0.19 | 0.029 | 0.47 | <0.001 |
| Grasela et al | -0.83 | <0.001 | 5.46 | <0.001 | 0.49 | <0.001 | -1.21 | <0.001 |

Figure S1 Overview of literature search

Figure S2. Goodness–Of-Fit plot of population predicted concentrations (ug/mL) versus observed concentrations (ug/mL) of phenobarbital for the external validation dataset and population pharmacokinetic models. The red line represents the identity line (y=x).

Figure S2. Normalized prediction distribution error (NPDE) plots of external validation data set for the investigated models, Moffett *et al*. (A), Yukawa *et al*. (B), Voller *et al*. (C), Vucicevic *et al*. (D), Grasela *et al*. (E): Quantile-quantile (q-q) plot of NPDE versus the expected standard normal distribution (upper left). Histogram of npde with the density of the standard normal distribution (upper right). The blue prediction intervals are obtained from the theoretical normal distribution. Scatterplot of the npde versus time after first dose in hour (lower left). The pink areas are the prediction interval for the median, while the blue areas show the prediction areas for the boundaries of the 95% prediction intervals. Scatterplot of npde versus predicted concentrations in ug/mL (lower right).

Figure S4. Box plots of individual prediction errors of population pharmacokinetic models by Vucicevic *et al*. with Bayesian forecasting in different number of prior measurements; 0, 1 2, 3 and all the observed measurements. Thick solid line represents the mean error of 0 and dotdashed line corresponds to the range of -30% and 30%.

Figure S5. Box plots(upper) and diagnostic plots(bottom) of prediction errors without or with incorporation of size or maturation function of each candidate models. Box plots were plotted from the predicted concentrations with estimated thetas with the external dataset, without size or maturation function, with size function, with maturation function, or with both size and maturation function, respectively. Thick solid line represents the mean error of 0 and dotdashed line corresponds to the range of -30% and 30%. In the goodness-of-fit plots, predicted concentration versus observed measurement were shown before (left) and after (right) incorporation of size or/and maturation function into the model. The red line represents the identity line.

